# Interthalamic adhesion alterations in multiple sclerosis: associations with thalamic damage and cognition

**DOI:** 10.64898/2026.01.31.26345261

**Authors:** Arthur Fournet, Fanny Munsch, Ismail Koubiyr, Julie P. Vidal, Sergio Morell-Ortega, Aurore Saubusse, Julie Charré-Morin, Katy K. Lataste, Audrey Lavielle, Zhang Bei, Pierrick Coupé, Emmanuel J. Barbeau, Vinod J. Kumar, Michael Hornberger, Vincent Dousset, Bruno Brochet, José V. Manjón, Aurélie Ruet, Thomas Tourdias

## Abstract

**Background / Objectives:** We investigated whether the interthalamic adhesion (IA), a midline structure connecting the thalami, is altered in MS and associated with thalamic damages and cognition.

**Methods:** We prospectively included 32 clinically isolated syndrome/early MS, 31 RRMS, 31 PPMS patients, and 103 matched controls. All underwent anatomical 3T MRI and completed a comprehensive cognitive battery. IA presence, subtype, and volume were assessed by two blinded readers. Thalamic nuclei and other brain structures were segmented automatically. We compared IA subtypes/volumes across groups, analyzed their predictors and explored cognitive associations with multivariate regressions.

**Results:** IA prevalence did not differ between MS and controls (81.9% vs 74.7%). MS patients showed a shift toward a short IA subtype and reduced IA volume (mean [SD], 146.8 [117.9] vs 230.2 [138.2] mm³; p<0.0001), worsening across phenotypes. Reduced IA volume was independently associated with medial and posterior thalamic nuclei volumes, but not with white matter lesion load or global atrophy. Among cognitive domains, smaller IA volume was independently associated only with executive dysfunction (OR = 0.89 [0.77–0.99], *p* = 0.021).

**Conclusion:** IA volume reduction in MS reflects vulnerability of adjacent thalamic nuclei and is associated with executive dysfunction, supporting IA as a marker of thalamic neurodegeneration.

**Trial Registration:** MICROSEP: NCT03692975; AUBACOG: NCT03768648; PROCOG: NCT03455582.

## Introduction

Multiple sclerosis (MS) involves acute inflammatory attacks but also chronic neurodegenerative processes that remain hard to specifically control (1). The latter drives relentless accumulation of disability, including cognitive decline and fatigue that can be present from the earliest disease stages (2) independently of overt inflammatory activity (3). Better imaging biomarkers that specifically capture neurodegeneration could support individualized therapeutic strategies and provide endpoints for future neuroprotective trials.

Gray matter atrophy reflects neuronal loss and is a robust biomarker of neurodegeneration (4). Beyond global measures, regional atrophy offers greater sensitivity for detecting early changes, particularly within selectively vulnerable structures (5). The thalamus has been identified as one of the earliest and most affected regions (6), with thalamic atrophy correlating strongly with clinical outcomes (7). Recent work has further emphasized differential vulnerability among thalamic nuclei (8, 9), with an “ependymal-in” gradient of damage linked to meningeal inflammation (10), preferentially affecting nuclei adjacent to the third ventricle. However, the specific involvement of medial thalamic nuclei, and their contribution to symptoms such as cognitive impairment, remains incompletely understood.

The medial thalami are interconnected by the interthalamic adhesion (IA), a small bridge of tissue traversing the third ventricle (11, 12). Long regarded as a vestigial structure, the IA has more recently been shown to contain fibers connecting both thalami as well as projecting to limbic and frontal networks (13, 14), suggesting a potential role in cognitive functions (15, 16). The IA is absent in 10 to 30% of the healthy population (11), which has been associated with increased vulnerability to some neuropsychiatric conditions such as schizophrenia or psychosis (17, 18), which is still debated (12). Direct alterations of the IA in neurological diseases have been scarcely investigated (19), and to date, no dedicated studies have explored its integrity in MS.

Given its strategic location bridging the medial thalamic nuclei, which are among the most vulnerable gray matter structures in MS (8, 9), we hypothesized that the IA may undergo variable disease-related morphological alterations across disease phenotypes. We further postulated that such changes could contribute to cognitive impairment in MS regarding IA connectivity with limbic and frontal networks (13, 14). Therefore, this study aimed to investigate IA alterations in MS, with the goal of advancing understanding of thalamus-related pathology and identifying a potential novel imaging-biomarker.

## Materials and methods

### Population

We prospectively included patients with MS and age- and sex-matched healthy controls (HCs) from three single-center cohorts investigating cognitive impairment across MS stages: MICROSEP (NCT03692975), AUBACOG (NCT03768648), and PROCOG (NCT03455582). All protocols were IRB-approved, and written informed consent was obtained.

MICROSEP included participants with clinically isolated syndrome (CIS) or early MS enrolled within six months of a first demyelinating event, requiring ≥2 clinically silent T2/FLAIR lesions (≥3 mm), including ≥1 ovoid or periventricular cerebral lesion. Early MS was defined by the 2017 McDonald criteria (20); others were classified as CIS. AUBACOG and PROCOG included patients with relapsing-remitting MS (RRMS) and primary progressive MS (PPMS) respectively, diagnosed per the 2017 McDonald criteria (20).

All participants completed identical neuropsychological assessments and MRI protocol. HCs recruited across studies were pooled into a single control group for the present analysis.

### Clinical evaluation and neuropsychological tests

The Expanded Disability Status Scale (EDSS) was administered by trained neurologists and a comprehensive neuropsychological battery was administered by two experienced neuropsychologists.

We evaluated information processing speed, attention, working memory, verbal memory, visuospatial memory and executive functions. Test performances were converted to HC-referenced Z-scores with impairment defined as Z-score <-1.5, in accordance with established thresholds (21). Details of the tests and number of abnormal tests required to consider a domain as significantly impaired are provided in **supplementary-material**.

### MRI acquisitions

All participants underwent brain MRI on a 3T scanner (Vantage Galan 3T/ZGO; Canon Medical Systems) that included a 3D gradient-echo T1-weighted, a 3D fluid-attenuated inversion recovery (FLAIR), and a 3D white matter-nulled magnetization-prepared rapid gradient echo (WMn-MPRAGE). Details of MR parameters are in **supplementary-material**.

We had previously optimized the WMn-MPRAGE to enhance visualization of internal thalamic anatomy and MS-related thalamic lesions (8, 22, 23). This sequence was reconstructed with a standard filter but also post-processed using a deep learning-based denoising technique called AiCE (Advanced Intelligent Clear-IQ Engine) (24). In addition, a third reconstruction of the WMn-MPRAGE was generated using a combined denoising (identical to the AiCE) and threefold k-space up-sampling method termed PIQE (Precise IQ Engine) (25), resulting in an apparent resolution of 0.5×0.3×0.3 mm^3^.

### Image analyses

#### Manual Identification of the IA

The 3D-T1 and the 3D-WMn-MPRAGE images with PIQE were cropped around the thalamus to minimize group-identifying cues and jointly presented in random order to two blinded readers (AF and TT) who conducted independent evaluations using established methodology (19), followed by a consensus review for final decision.

Using the combined T1 and WMn-MPRAGE, IA was considered present if a structure connecting the bilateral thalami between the anterior and posterior commissures was visible on ≥ 1 axial slice and confirmed in orthogonal planes. IA subtypes were classified as before (19) as short, broad, bilobar, double, or rudimental. Specifically, IA was defined as broad if extending over ≥ 1/3 of thalamic length or height. When thalami were in direct contact across multiple slices due to a narrow third ventricle, IA assessment was not possible; a situation labeled “kissing thalami”(13).

Then, IA was manually segmented on consecutive sagittal slices using 3D Slicer (v5.4.0) to measure volume and maximal height and length.

The WMn was also used to identify thalamic lesions as described before (23) and their proximity with IA.

#### Comparison of WMn reconstructions

To assess how different image reconstructions might influence IA visibility, one reader (TT) manually traced a line throughout the IA to the adjacent third ventricle cerebrospinal fluid (CSF) on axial slices of the raw WMn (standard filter), as well as on the AiCE and PIQE reconstructions. These lines were used to extract signal intensity profiles across the IA-CSF transitions which were compared between the reconstruction methods. The anterior transition was a signal decrease from hypersignal of CSF to low signal of IA, while the posterior transition from IA to CSF was opposite. Therefore, we analyzed the maximum negative and positive slopes computed through the first derivative of each profile with respect to distance to quantify the sharpness of these transitions.

#### Volumetry of other brain structures

White matter MS lesions were automatically segmented from 3D-FLAIR and 3D-T1 using DeepLesionBrain (26) followed by a visual quality-control.

Total brain, white matter, gray matter, and CSF volumes were derived from 3D-T1 using AssemblyNet (27).

Thalamic nuclei were segmented on 3D-WMn images using the THOMAS multi-atlas pipeline (28) and grouped into four anatomically and functionally defined regions (anterior, lateral, medial, and posterior thalamic groups) (29).

All volumes were normalized using the affine MNI-based volumetric scaling factor.

#### Automatic Delineation of the IA

As the IA is not included in any currently available automatic brain structure segmentation tools, we leveraged the manually defined 3D IA segmentations to train a deep convolutional neural network (CNN). Details of the CNN architecture are in **supplementary-materials**.

A fully independent validation set of 30 other healthy participants with both 3D-T1 and 3D-WMn acquisitions was used specifically for external testing. These cases were reviewed to assess IA presence and manually delineate the structure as described in the training phase. The CNN-derived segmentations were then compared to this “ground truth” using the Dice similarity coefficient, Pearson correlation, and the 95th percentile Hausdorff distance.

### Statistical analyses

Signal intensity profiles across the standard WMn, AiCE, and PIQE reconstructions were compared using repeated-measures ANOVA with Dunnett’s post hoc tests.

Inter-reader reproducibility was assessed using Cohen’s kappa coefficient for IA subtype classification and intraclass correlation coefficient (ICC) for quantitative measurements.

IA subtype distributions were compared between groups using chi-square tests, and quantitative IA measures were compared using one-way ANOVA with Dunnett’s multiple comparison tests.

In patients and HCs with IA presence, factors associated with IA volume were examined using multiple linear regressions with the following potential predictors: age, sex, lesion load, CSF volume, total brain volume, whole thalamus volume, and the volumes of thalamic nuclei groups. Multicollinearity was assessed using variance inflation factors (VIF).

Finally, in an exploratory analysis, IA volumes were compared between impaired and preserved patients across the six cognitive domains using Mann-Whitney tests. For cognitive domains showing group differences, a multivariable logistic regression was performed with domain-specific impairment as the dependent variable.

All statistical analyses were conducted using GraphPad Prism (10.4.1).

## Results

### Patient characteristics

We included 197 participants, comprising 103 HCs and 94 patients with a balanced distribution of CIS/early MS (n = 32), RRMS (n = 31), and PPMS (n = 31). Groups were matched for age, sex, and educational level, with the exception of the PPMS group which was significantly older. **Table 1** summarizes the clinical characteristics.

**Table 1:** Demographic characteristics.

|  | **Healthy controls**<br>(n=103) | **Patients**<br>(n=94) | **CIS/early MS**<br>(n=32) | **RRMS**<br>(n=31) | **PPMS**<br>(n=31) |
| --- | --- | --- | --- | --- | --- |
| **DEMOGRAPHICS** |  |  |  |  |  |
| Age (years), mean (SD) | 39.2 (9.3) | 43.3 (11.0) | 34.9 (9.3) | 43.5 (9.3) | 51.9 (6.9) |
| Sex (female), n (%) | 70 (67.9) | 65 (69.1) | 24 (75.0) | 20 (64.5) | 21 (67.7) |
| Disease duration (years), mean (SD) | NA | 4.94 (6.16) | 0.36 (0.13) | 10.3 (7.2) | 4.20 (3.10) |
| Educational level (Baccalaureate), n (%) | 78 (75.7) | 69 (73.4) | 27 (84.3) | 24 (77.4) | 18 (58.1) |
| EDSS (score), median (Q1, Q3) | NA | 2 (1, 3) | 1.5 (1, 2.5) | 1.5 (0, 2.5) | 3.5 (2.5, 4.5) |
| Beck depression inventory (score), median (Q1, Q3) | 4 (2, 10) | 10.5 (5, 16) | 8 (3.5, 15) | 8 (4.7, 14.5) | 13.5 (9, 20) |
| **COGNITIVE EVALUATIONS** |  |  |  |  |  |
| Information processing speed (% impaired) | 6.0 | 30.2 | 18.7 | 32.3 | 43.5 |
| Attention (% impaired) | 22.0 | 31.4 | 28.1 | 25.8 | 43.5 |
| Working memory (% impaired) | 15.0 | 32.6 | 25 | 35.5 | 39.1 |
| Verbal memory (% impaired) | 12.0 | 25.6 | 12.5 | 29.0 | 39.1 |
| Visio-spatial memory (% impaired) | 9.0 | 16.3 | 9.4 | 19.4 | 21.7 |
| Executive functions (% impaired) | 7.0 | 15.1 | 6.3 | 19.4 | 21.7 |

CIS: clinically isolated syndrome; EDSS: expanded disability status scale; MS: multiple sclerosis; NA: not available; PPMS: primary progressive multiple sclerosis; RRMS: relapsing remitting multiple sclerosis;

### WMn images with super-resolution ensure reliable IA assessment

We supplemented the standard WMn reconstruction with denoising (AiCE) and up-sampling (PIQE) algorithms. Signal intensity profiles showed significantly steeper transitions at the IA-CSF interface with the denoising and super-resolution that progressively enhanced image sharpness, an objective indicator of improved image quality (**Figure 1**).

**Figure 1.**
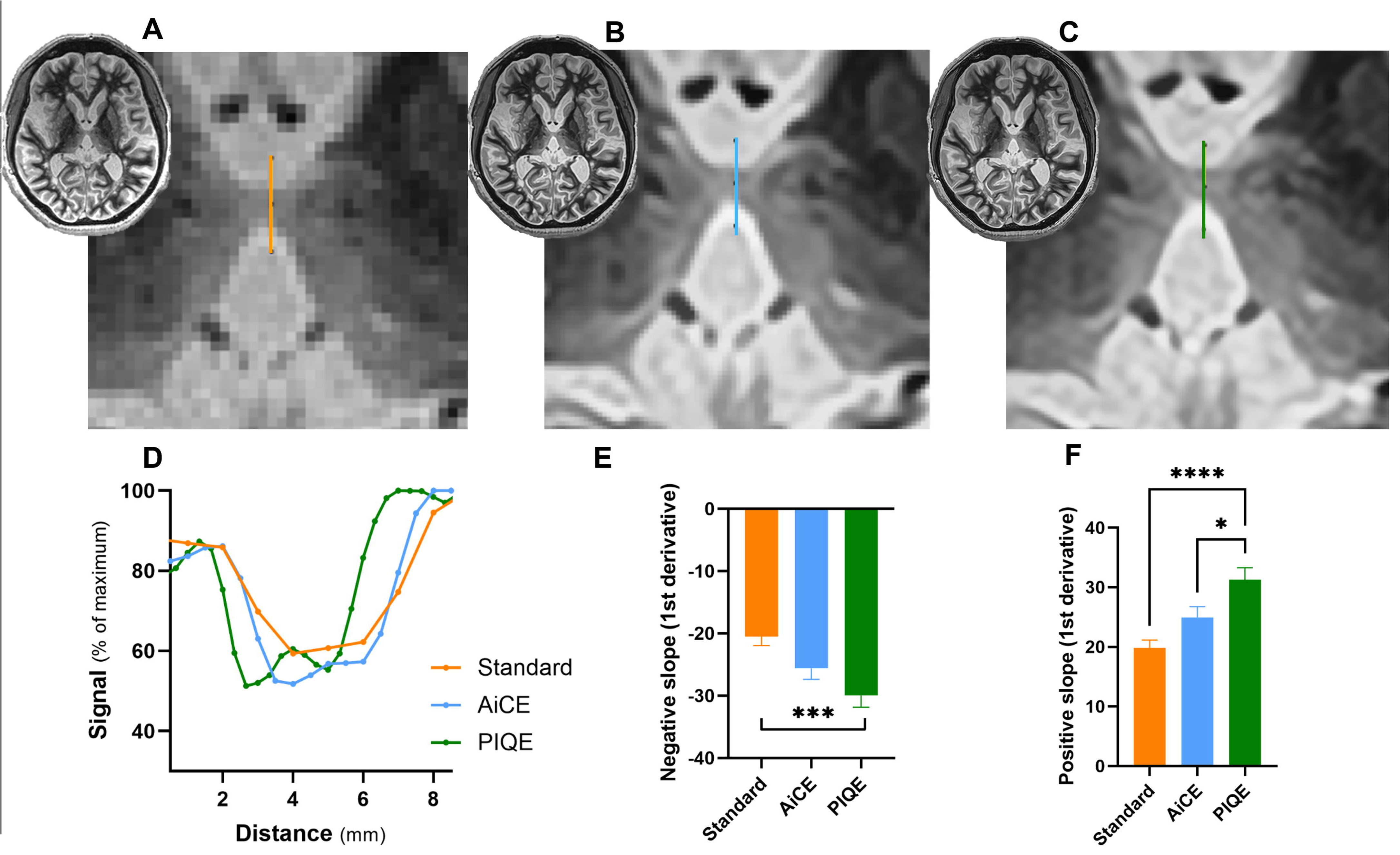
Effect of denoising and super-resolution on white matter-nulled (WMn) images for visualization of the interthalamic adhesion (IA) Representative aspect of IA on WMn images reconstructed using the standard method (**A**), a deep learning-based denoising algorithm (AiCE) (**B**), and the same algorithm combined with an upsampling technique (PIQE) (**C**). Representative signal intensity profiles along lines traversing the IA (**D)**, and positive and negative slopes at the IA/CSF interfaces (**E and F**). Quantitative analysis of the positive and negative slopes of intensity transitions across the entire population shows significantly steeper slopes with PIQE, reflecting enhanced edge definition. Data are presented as mean ± SEM. Statistical significance from Dunnett’s multiple comparisons test following one-way ANOVA is indicated as follows: *p = 0.03; ***p = 0.0007; ****p < 0.0001. AiCE, Advanced Intelligent Clear-IQ Engine; PIQE, Precise IQ Engine

Using these enhanced images, the two readers achieved substantial agreement in identifying the presence of the IA and classifying it into subtypes, with a Cohen’s kappa of 0.71 (95% CI, 0.63–0.80) (**Figure 2A**). Disagreements could occur with the “kissing thalami” configuration, which could be interpreted for an absent IA due to poor visibility.

**Figure 2.**
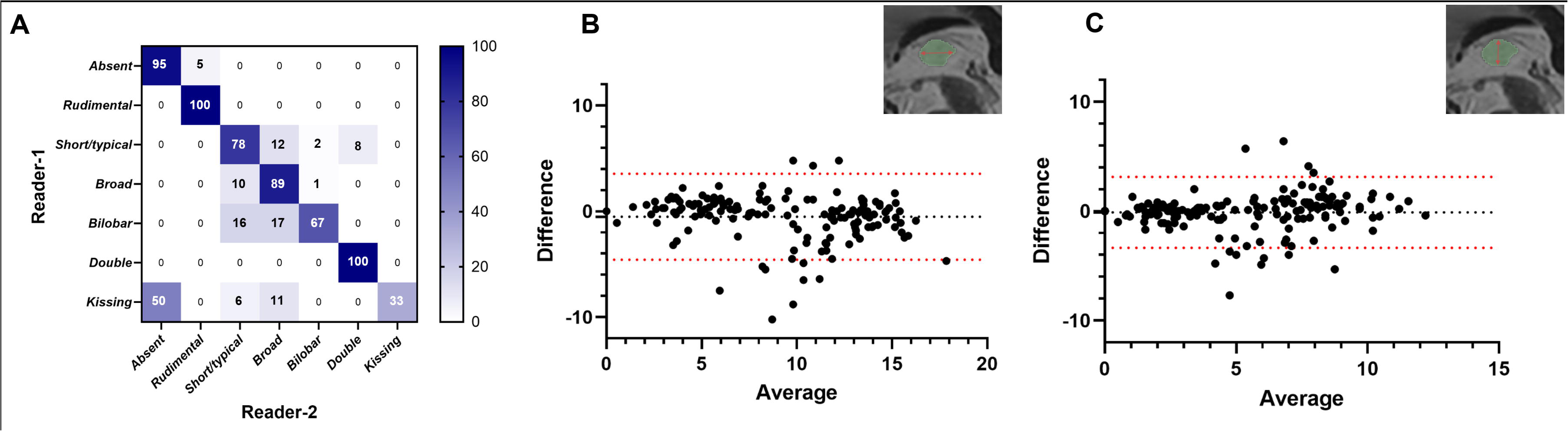
Reproducibility of IA Evaluations. Correlation matrix showing the percentage of agreement between the two independent readers in classifying IA subtypes (**A**). Bland–Altman plots illustrating inter-reader agreement for IA length and height, by showing the difference between measurements as a function of their mean (**B and C**). The black dotted line indicates the mean bias, and the red dotted lines represent the 95% limits of agreement.

Despite the small size of the structure, the quantitative measurements showed good-to-excellent inter-reader reproducibility. ICC were 0.91 (95% CI, 0.84-0.87) for IA length and 0.88 (95% CI, 0.83-0.85) for IA height (**Figure 2B–C**), with mean absolute biases of –0.50 mm and –0.11 mm, respectively.

### IA can be automatically delineated with a deep-learning-based algorithm

Although manual IA identification and delineation yielded good reproducibility, the process was time-consuming and labor-intensive. To address this, we used the 197 manual delineations to train a deep CNN.

External validation in 30 unseen new participants showed perfect detection of IA absence (8/8) and strong agreement with manual segmentations when present (r = 0.89, *p* < 0.00001), with median Dice of 0.64 (IQR, 0.58–0.68) and median 95th percentile Hausdorff distance of 1.9 mm (IQR, 1.1–2.4) (**Supplementary-Figure 1**).

### Variation of IA anatomy is encountered in MS patients

IA prevalence was similar in HCs and MS patients (74.7% vs. 81.9%, *p* = 0.23, Chi² test), with remaining cases showing no visible IA or a non-assessable “kissing thalami” configuration (**Table-2**).

**Table 2:** Imaging characteristics.

|  | Healthy controls<br>(n=103) | Patients<br>(n=94) | CIS/early MS<br>(n=32) | RRMS<br>(n=31) | PPMS<br>(n=31) |
| --- | --- | --- | --- | --- | --- |
| **INTER-THALAMIC ADHESION (IA)** |  |  |  |  |  |
| IA prevalence |  |  |  |  |  |
| Presence of IA, n (%) | 77 (74.8) | 77 (81.9) | 25 (78.1) | 23 (74.2) | 29 (93.5) |
| Absence of IA, n (%) | 14 (13.6) | 14 (14.9) | 5 (15.6) | 7 (22.6) | 2 (6.5) |
| Kissing thalami, n (%) | 12 (11.6) | 3 (3.2) | 2 (6.2) | 1 (3.2) | 0 (0) |
| IA subtype |  |  |  |  |  |
| Rudimental, n (%) | 2 (1.9) | 1 (1.1) | 1 (3.1) | 0 (0) | 0 (0) |
| Short, n (%) | 18 (17.5) | 37 (39.4) | 5 (15.6) | 13 (41.9) | 19 (61.3) |
| Broad, n (%) | 53 (51.4) | 30 (31.9) | 18 (56.2) | 7 (22.6) | 5 (16.1) |
| Bilobar, n (%) | 3 (2.9) | 4 (4.2) | 1 (3.1) | 0 (0) | 3 (9.7) |
| Double, n (%) | 1 (0.9) | 5 (5.3) | 0 (0) | 3 (9.7) | 2 (6.4) |
| IA volumetry |  |  |  |  |  |
| IA length (mm), mean (SD) | 10.78 (3.93) | 7.76 (3.96) | 10.02 (4.45) | 7.09 (2.96) | 6.35 (3.41) |
| IA height (mm), mean (SD) | 6.50 (2.67) | 4.30 (2.72) | 5.79 (2.77) | 3.76 (2.25) | 3.44 (2.56) |
| IA volume (mm<sup>3</sup>), mean (SD) | 230.2 (138.2) | 146.8 (117.9) | 210.4 (126.7) | 135.0 (112.9) | 101.2 (90.03) |
| **OTHER IMAGING FEATURES** |  |  |  |  |  |
| Lesion load (cm<sup>3</sup>), mean (SD) | NA | 8.07 (14.9) | 2.67 (2.32) | 8.10 (6.38) | 13.8 (24.1) |
| Total brain volume (cm<sup>3</sup>), mean (SD) | 1609 (43.44) | 1567 (70.97) | 1610 (48.84) | 1570 (53.05) | 1520 (77.18) |
| CSF volume (cm<sup>3</sup>), mean (SD) | 228.1 (44.60) | 263.1 (62.77) | 221.6 (35.93) | 268.2 (60.12) | 300.9 (62.55) |
| White matter volume (cm<sup>3</sup>), mean (SD) | 608.2 (26.42) | 584 (42.49) | 608 (32.05) | 584.1 (34.29) | 559.1 (45.64) |
| Gray matter volume (cm<sup>3</sup>), mean (SD) | 1001 (30.57) | 983.2 (34.79) | 1002 (27.33) | 986.3 (27.75) | 960.7 (35.78) |
| Total thalamic volume (cm<sup>3</sup>), mean (SD) | 7.48 (0.53) | 6.65 (1.03) | 7.28 (0.7) | 6.51 (0.89) | 6.12 (1.11) |
| Anterior thalamic volume (cm<sup>3</sup>), mean (SD) | 0.164 (0.003) | 0.141 (0.049) | 0.168 (0.005) | 0.131 (0.046) | 0.123 (0.039) |
| Lateral thalamic volume (cm<sup>3</sup>), mean (SD) | 1.994 (0.152) | 1.794 (0.270) | 1.946 (0.199) | 1.737 (0.236) | 1.688 (0.296) |
| Posterior thalamic volume (cm<sup>3</sup>), mean (SD) | 2.082 (0.193) | 1.761 (0.367) | 1.972 (0.278) | 1.707 (0.327) | 1.591 (0.384) |
| Medial thalamic volume (cm<sup>3</sup>), mean (SD) | 1.092 (0.084) | 0.981 (0.148) | 1.067 (0.102) | 0.965 (0.143) | 0.906 (0.151) |

CIS: clinically isolated syndrome; CSF: cerebrospinal fluid; IA: inter-thalamic adhesion; MS: multiple sclerosis; NA: not available; PPMS: primary progressive multiple sclerosis; RRMS: relapsing remitting multiple sclerosis;

The five anatomical IA subtypes are illustrated in **Supplementary-Figure 2**. On WMn images, the IA did not appear completely nulled, in contrast to other commissures such as the anterior commissure or the corpus callosum suggesting that this is not a pure white matter structure (**Supplementary-Figure 3**).

In HCs, the IA most commonly appeared as the broad subtype (51.4%). In contrast, there was a progressive and statistically significant shift toward the short subtype across MS phenotypes; 15.6% in CIS/early MS, 41.9% in RRMS, and 61.3% in PPMS (*p* < 0.001, Chi² test, **Table-2**).

### IA is altered in MS patients in relation with damages to the thalamic nuclei close to the third ventricle

IA volume was significantly reduced in MS patients compared to controls (146.8 mm³ vs. 230.2 mm³; *p* < 0.0001), with a progressive decline across disease phenotype, from CIS/early MS to RRMS to PPMS (*p* < 0.001, one-way ANOVA) (**Figure 3 and Table 2**). Similar patterns were observed when analyzing IA length and height (**Supplementary-Figure 4**).

**Figure 3.**
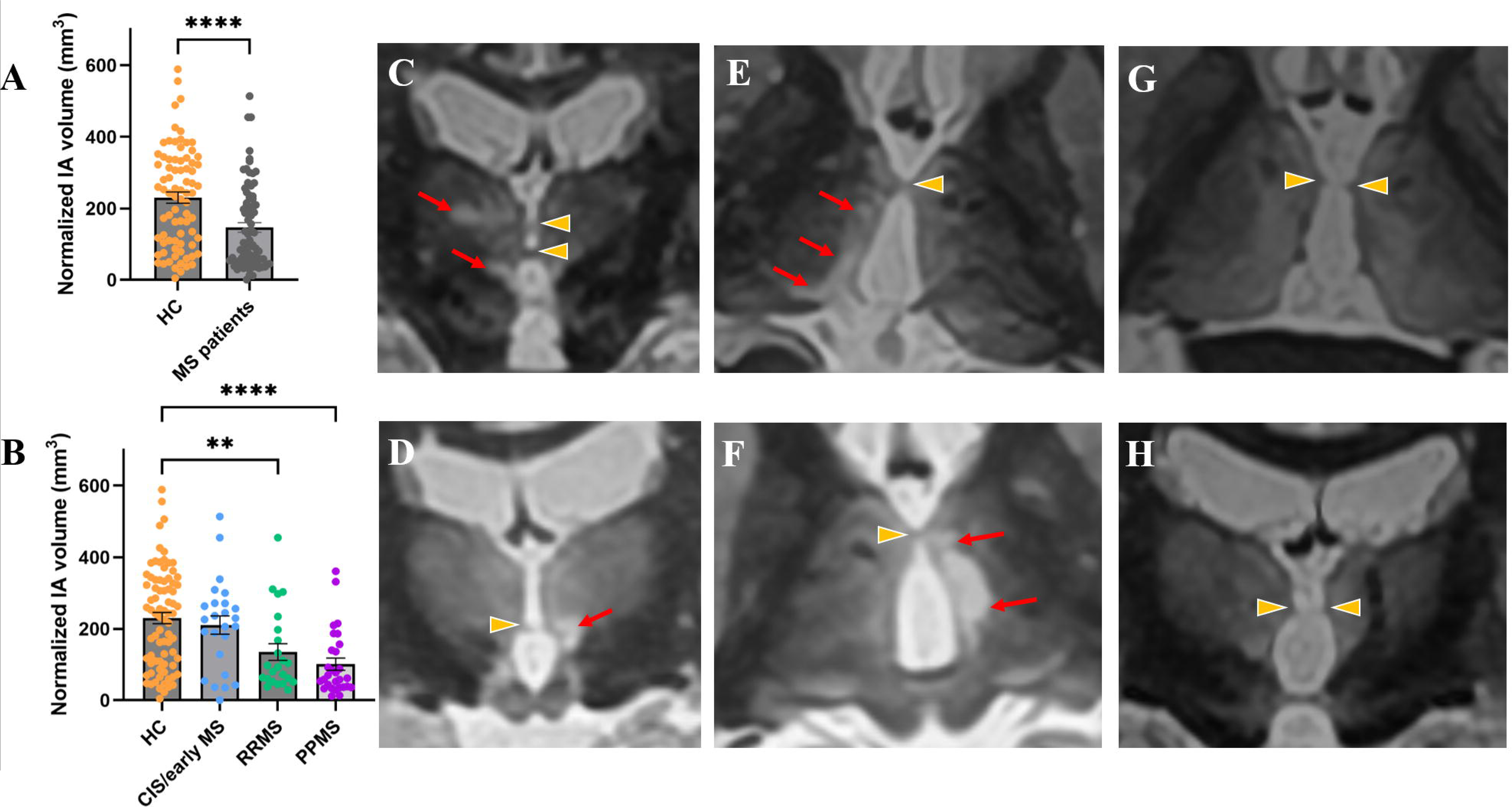
Alteration of IA in MS patients. IA volumes in healthy controls (HC) and patients with MS (**A-B**). Data are presented as mean ± SEM. Statistical significance from Dunnett’s multiple comparisons test following one-way ANOVA is indicated as follows: **p = 0.0048; **** p < 0.0001. Illustrative WMn images (coronal plane for C, D, H and axial plane for E, F, G) showing focal thalamic lesions mainly appearing as band-like structures adjacent to the third ventricle (red arrows), sometimes in close proximity to a small IA (yellow arrowheads) (**C-F**). Illustrative examples of virtually absent IA but associated with remnant-like structures at the ependymal surface (**G-H**). CIS, clinically isolated syndrome, RRMS, relapsing remitting multiple sclerosis, PPMS, primary progressive multiple sclerosis.

**Figure 4.**
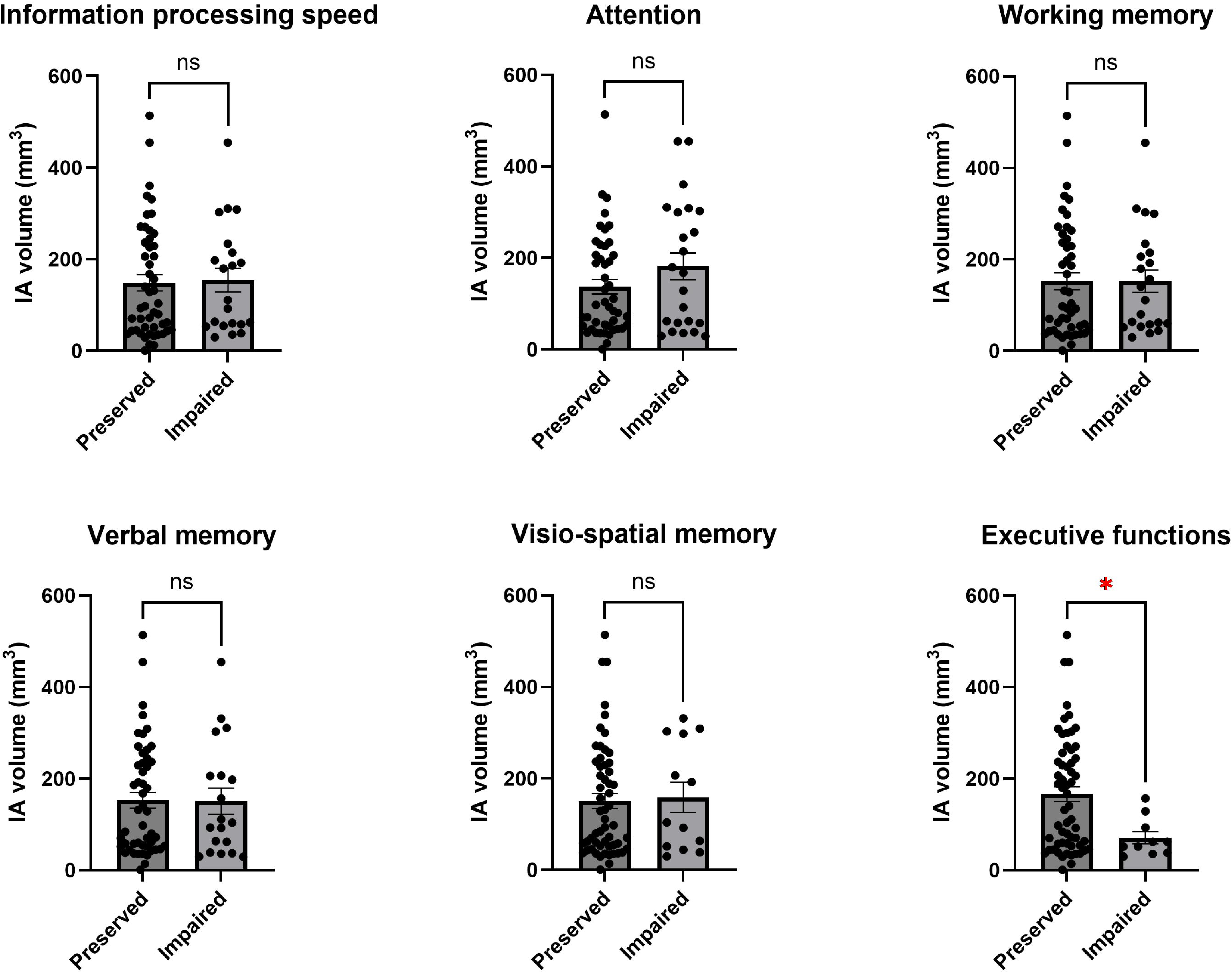
Relation between IA and cognitive status. IA volumes are presented according to cognitive status in each of the six domains. Statistical significance for Mann-Whitney tests as follows: *p = 0.02.

WMn revealed thalamic lesions that predominantly localized along the walls of the third ventricle. In some individuals, these band-like lesions appeared to contact a particularly small IA (**Figure 3C-F**). In other cases, the absence of the central portion of the IA was associated with remnant-like structures at the ependymal surface, suggestive of secondary atrophy rather than a primary anatomical absence (**Figure 3G-H**).

To better understand the factors influencing IA volume, we conducted multiple linear regression analyses. Results showed that lower IA volume was significantly and independently associated with male sex and lower whole thalamus volume, but not with age, lesion load, or global atrophy markers such as brain or CSF volumes (Model 1, **Table 3**).

**Table 3:** Linear regression models for association with inter-thalamic adhesion volume.

| Independent variables | Estimate | 95% CI | P value |
| --- | --- | --- | --- |
| **MODEL 1** |  |  |  |
| Age | -0.6523 | -3.159 to 1.855 | 0.6078 |
| Sex [female] | 72.76 | 28.30 to 117.2 | **0.0015\*\*** |
| Lesion load | 0.001008 | -0.001321 to 0.003336 | 0.3936 |
| CSF volume | -0.1413 | -0.8108 to 0.5282 | 0.6771 |
| Brain Volume | 0.2066 | -0.4457 to 0.8590 | 0.5321 |
| Whole Thalamus Volume | 0.05054 | 0.01550 to 0.08558 | **0.005\*\*** |
| **MODEL 2** |  |  |  |
| Age | -1.061 | -3.539 to 1.416 | 0.3984 |
| Sex [female] | 69.36 | 24.94 to 113.8 | **0.0024\*\*** |
| Lesion load | 0.0005874 | -0.001646 to 0.002821 | 0.6038 |
| CSF volume | -0.159 | -0.8287 to 0.5107 | 0.6396 |
| Brain Volume | 0.1944 | -0.4637 to 0.8525 | 0.5601 |
| Medial thalamic group Volume | 0.311 | 0.08860 to 0.5333 | **0.0065\*\*** |
| **MODEL 3** |  |  |  |
| Age | -0.6501 | -3.169 to 1.869 | 0.6107 |
| Sex [female] | 71.69 | 27.15 to 116.2 | **0.0018\*\*** |
| Lesion load | 0.0007877 | -0.001504 to 0.003080 | 0.4979 |
| CSF volume | -0.1242 | -0.7972 to 0.5489 | 0.7158 |
| Brain Volume | 0.2828 | -0.3591 to 0.9248 | 0.3851 |
| Posterior thalamic group Volume | 0.1228 | 0.03281 to 0.2128 | **0.0078\*** |
| **MODEL 4** |  |  |  |
| Age | -0.891 | -3.443 to 1.661 | 0.4911 |
| Sex [female] | 71.24 | 25.96 to 116.5 | **0.0023\*\*** |
| Lesion load | 0.0004204 | -0.001920 to 0.002761 | 0.723 |
| CSF volume | -0.2151 | -0.8939 to 0.4637 | 0.532 |
| Brain Volume | 0.3656 | -0.2832 to 1.014 | 0.2671 |
| Lateral thalamic group Volume | 0.1028 | -0.01340 to 0.2191 | 0.0825 |

Because of collinearity (VIF > 10), whole thalamus volume and individual nuclei group volumes could not be included in the same model. Replacing the total thalamus volume with nuclei group volumes showed that IA volume remained independently associated with sex and medial or posterior, but not lateral, thalamic groups (Models 2-4, **Table 3**), suggesting IA as a proxy for alterations in thalamic structures adjacent to CSF.

### Relation with cognitive status

Patients with MS exhibited lower cognitive Z-scores than HCs (**Supplementary-Figure 5**) resulting in a higher proportion meeting the threshold for clinically significant impairment across all cognitive domains from CIS to RRMS to PPMS (**Table 1**).

When stratifying the entire group of MS patients by cognitive status, IA volume was significantly reduced only in those with marked executive dysfunction (*p* = 0.02). Logistic regression showed that a lower IA volume was significantly associated with higher probability of impairment in executive functions independently from age, gender, educational level, lesion load, whole brain volume and whole thalamus volume (OR_(10mm3)_ = 0.89 [0.77–0.99], *p* = 0.021).

## Discussion

Through optimized image acquisition and analysis, we provide the first evidence of morphological alterations of the IA in MS, with progressive changes in shape and volume across disease phenotypes. These alterations appear linked to the vulnerability of thalamic nuclei adjacent to the third ventricle and may contribute to impairments in specific cognitive domains.

We based IA identification and delineation on 3D-T1-weithed MR images as in prior studies (13, 15, 16, 19), but also leveraged a white matter-nulled (WMn) version of MPRAGE sequence, specifically tuned for this particular application with denoising and super-resolution steps to enhance further detectability and sharpness. Because WMn imaging simultaneously reveals thalamic nuclei (22), otherwise inconspicuous MS lesions within these nuclei (23), and the IA itself, it provided the unique opportunity to examine whether IA alterations are linked to nearby thalamic lesions or to structural changes in nuclei bordering the third ventricle.

Using this approach, we found IA prevalence ranging from ∼75% in controls to ∼82% in MS patients, in line with previous reports (11, 12, 15, 19), though variability likely reflects methodological and population-specific differences. Beyond prevalence, WMn imaging provided novel insights into IA composition, which has long been debated between a gray matter-like structure with neuronal cell bodies (30) or rather a white matter-like structure with fibers (14, 31). IA aspect on WMn that is not completely nulled, supports the view that IA is a mixed structure containing both central gray matter neurons and interconnecting white matter fibers, with regional differences in cellular density between inferior and superior segments.

Previous studies reported higher IA absence rates in some psychiatric conditions (17, 18), but no volumetric analyses had been conducted in neurological diseases. In MS, we observed a similar absence rate to the general population but documented progressive IA volume loss from early disease to relapsing-remitting MS and into progressive stages. This atrophy was driven by thalamic rather than global brain volume loss and was specifically associated with nuclei bordering the third ventricle and with subpial band-like lesions. Taken together, these results question whether IA alterations could be related to meningeal inflammation, a mechanism increasingly implicated in thalamic neurodegeneration (9, 10, 32), and whether IA could be a candidate biomarker of this process.

We also provided first preliminary data suggesting a possible association between IA volume and executive function performance which further supports a functional role of IA in interhemispheric integration. Tractography studies have shown that IA connects the prefrontal cortex with the contralateral mediodorsal thalamus via anterior thalamic nuclei, the stria medullaris, and the habenular nucleus (13, 33) - pathways central to planning, inhibition, and cognitive flexibility. IA disruption may therefore contribute specifically to executive dysfunction. By contrast, other cognitive domains such as information processing speed or memory known to depend on diffuse white matter pathology or hippocampo-temporal circuits respectively (2) might be less dependent of IA-associated network. These data align with the emerging view that IA can contribute to cognition (15, 16, 19) and raise the possibility of compensatory pathways in individuals without this structure.

In parallel, we capitalized on careful multi-reader manual assessment of IA to develop and release a deep learning-based tool for automatic IA delineation, a structure that is not included in any of the currently available automated volumetric tools. This solution offers a foundation for larger studies to consolidate IA role as a potential imaging marker of neurodegeneration in MS. The pipeline was originally trained with T1 and WMn images, but can also be run from standard T1 only with an intermediate step of WMn synthesis (34, 35), thus broadening applicability to existing cohorts.

We have to acknowledge limitations to contextualize the interpretation of our findings. First, the IA’s small size makes it vulnerable to partial volume effects with adjacent CSF and thalamus, which we did not explicitly correct. Second, while IA damage may be driven by direct alteration associated with thalamic damages, indirect disconnection effects from white matter lesions also warrant exploration using advanced tractography. Diffusion tensor and functional MRI approaches, adapted to the IA’s small scale, will be needed in future work. Third, the clinical correlations should be viewed as exploratory regarding the small sample size that limits capacities to fully control for all potential confounders. Finally, the cross-sectional design precludes conclusions on prognostic value. Longitudinal studies will be essential to clarify the trajectory of IA atrophy, its mechanisms, and its impact on future clinical outcomes.

In conclusion, we report IA morphological alterations associated with MS severity that reflect vulnerability of periventricular thalamic nuclei to neurodegenerative processes. The association with executive dysfunction challenges the view of IA as vestigial, instead suggesting a role as an indirect marker of interhemispheric connectivity and thalamo-prefrontal networks. This opens perspectives for monitoring IA and evaluating its potential as a marker of neurodegeneration and therapeutic response in MS which could be facilitated by the release of a new automatic delineation tool.

## Statements and declarations

### Ethical considerations

MICROSEP, AUBACOG, and PROCOG (NCT03455582) have been approved by the institutional review boards under the following approval numbers: MICROSEP (#18.021, CPP Ile de France V), AUBACOG (#18 78, CPP Sud Méditerranée I), PROCOG (#2017-72, CPP Nord Ouest II).

### Consent to participate

All participants have provided written inform consent.

### Consent for publication

Not applicable

## Data availability

The data that supported the findings of this study are available from the corresponding author upon reasonable request.

## References

1. Lassmann H. Pathogenic Mechanisms Associated With Different Clinical Courses of Multiple Sclerosis. Front Immunol. 2018;9:3116.

2. Chiaravalloti ND, DeLuca J. Cognitive impairment in multiple sclerosis. Lancet Neurol. 2008;7(12):1139–51.

3. Fuchs TA, Schoonheim MM, Zivadinov R, Dwyer MG, Colato E, Weinstock Z, et al. Cognitive progression independent of relapse in multiple sclerosis. Mult Scler. 2024;30(11-12):1468–78.

4. Popescu V, Klaver R, Voorn P, Galis-de Graaf Y, Knol DL, Twisk JW, et al. What drives MRI-measured cortical atrophy in multiple sclerosis? Mult Scler. 2015;21(10):1280–90.

5. Haider L, Zrzavy T, Hametner S, Hoftberger R, Bagnato F, Grabner G, et al. The topograpy of demyelination and neurodegeneration in the multiple sclerosis brain. Brain. 2016;139(Pt 3):807–15.

6. Coupe P, Planche V, Mansencal B, Kamroui RA, Koubiyr I, Manjon JV, et al. Lifespan neurodegeneration of the human brain in multiple sclerosis. Hum Brain Mapp. 2023;44(17):5602–11.

7. Eshaghi A, Prados F, Brownlee WJ, Altmann DR, Tur C, Cardoso MJ, et al. Deep gray matter volume loss drives disability worsening in multiple sclerosis. Ann Neurol. 2018;83(2):210–22.

8. Blyau S, Koubiyr I, Saranathan M, Coupe P, Deloire M, Charre-Morin J, et al. Differential vulnerability of thalamic nuclei in multiple sclerosis. Mult Scler. 2022:13524585221114247.

9. Koubiyr I, Yamamoto T, Blyau S, Kamroui RA, Mansencal B, Planche V, et al. Vulnerability of Thalamic Nuclei at CSF Interface During the Entire Course of Multiple Sclerosis. Neurol Neuroimmunol Neuroinflamm. 2024;11(3):e200222.

10. Magliozzi R, Fadda G, Brown RA, Bar-Or A, Howell OW, Hametner S, et al. “Ependymal-in” Gradient of Thalamic Damage in Progressive Multiple Sclerosis. Ann Neurol. 2022.

11. Wong AK, Wolfson DI, Borghei A, Sani S. Prevalence of the interthalamic adhesion in the human brain: a review of literature. Brain Struct Funct. 2021;226(8):2481–7.

12. Abramek K, Sobczak F, Walocha J, Ghosh S, Patra A, Balawender K. Exploring the interthalamic adhesion: A comprehensive review of its morphology, neuroconnectivity and history of research. Ann Anat. 2026;263:152728.

13. Borghei A, Kapucu I, Dawe R, Kocak M, Sani S. Structural connectivity of the human massa intermedia: A probabilistic tractography study. Hum Brain Mapp. 2021;42(6):1794–804.

14. Sahin MH, Gungor A, Demirtas OK, Postuk C, Firat Z, Ekinci G, et al. Microsurgical and fiber tract anatomy of the interthalamic adhesion. J Neurosurg. 2023;139(5):1386–95.

15. Damle NR, Ikuta T, John M, Peters BD, DeRosse P, Malhotra AK, et al. Relationship among interthalamic adhesion size, thalamic anatomy and neuropsychological functions in healthy volunteers. Brain Struct Funct. 2017;222(5):2183–92.

16. Borghei A, Cothran T, Brahimaj B, Sani S. Role of massa intermedia in human neurocognitive processing. Brain Struct Funct. 2020;225(3):985–93.

17. Asghar A, Narayan RK, Kumar P, Ravi KS, Tubbs RS, Patra A, et al. Absence of the interthalamic adhesion (ITA) as a neuroanatomical association or risk factor for neuropsychiatric disorders: A systemic review and meta-analysis. Indian J Psychiatry. 2023;65(10):985–94.

18. Erbagci H, Yildirim H, Herken H, Gumusburun E. A magnetic resonance imaging study of the adhesio interthalamica in schizophrenia. Schizophr Res. 2002;55(1-2):89–92.

19. Vidal JP, Rachita K, Servais A, Peran P, Pariente J, Bonneville F, et al. Exploring the impact of the interthalamic adhesion on human cognition: insights from healthy subjects and thalamic stroke patients. J Neurol. 2024;271(9):5985–96.

20. Thompson AJ, Banwell BL, Barkhof F, Carroll WM, Coetzee T, Comi G, et al. Diagnosis of multiple sclerosis: 2017 revisions of the McDonald criteria. Lancet Neurol. 2018;17(2):162–73.

21. Benedict RH, Zivadinov R. Reliability and validity of neuropsychological screening and assessment strategies in MS. J Neurol. 2007;254 Suppl 2:II22–II5.

22. Tourdias T, Saranathan M, Levesque IR, Su J, Rutt BK. Visualization of intra-thalamic nuclei with optimized white-matter-nulled MPRAGE at 7T. Neuroimage. 2014;84:534–45.

23. Planche V, Su JH, Mournet S, Saranathan M, Dousset V, Han M, et al. White-matter-nulled MPRAGE at 7T reveals thalamic lesions and atrophy of specific thalamic nuclei in multiple sclerosis. Mult Scler. 2019:1352458519828297.

24. Kidoh M, Shinoda K, Kitajima M, Isogawa K, Nambu M, Uetani H, et al. Deep Learning Based Noise Reduction for Brain MR Imaging: Tests on Phantoms and Healthy Volunteers. Magn Reson Med Sci. 2019.

25. Matsuo K, Nakaura T, Morita K, Uetani H, Nagayama Y, Kidoh M, et al. Feasibility study of super-resolution deep learning-based reconstruction using k-space data in brain diffusion-weighted images. Neuroradiology. 2023;65(11):1619–29.

26. Kamraoui RA, Ta VT, Tourdias T, Mansencal B, Manjon JV, Coup P. DeepLesionBrain: Towards a broader deep-learning generalization for multiple sclerosis lesion segmentation. Med Image Anal. 2022;76:102312.

27. Coupe P, Mansencal B, Clement M, Giraud R, Denis de Senneville B, Ta VT, et al. AssemblyNet: A large ensemble of CNNs for 3D whole brain MRI segmentation. Neuroimage. 2020;219:117026.

28. Su JH, Thomas FT, Kasoff WS, Tourdias T, Choi EY, Rutt BK, et al. Thalamus Optimized Multi Atlas Segmentation (THOMAS): fast, fully automated segmentation of thalamic nuclei from structural MRI. Neuroimage. 2019;194:272–82.

29. Herrero MT, Barcia C, Navarro JM. Functional anatomy of thalamus and basal ganglia. Childs Nerv Syst. 2002;18(8):386–404.

30. Laslo P, Slobodan M, Nela P, Milos M, Rade P, Tatjana I. Specific circular organization of the neurons of human interthalamic adhesion and of periventricular thalamic region. Int J Neurosci. 2005;115(5):669–79.

31. Parra JED, Ripoll AP, Garcia JFV. Interthalamic adhesion in humans: a gray commissure? Anat Cell Biol. 2022;55(1):109–12.

32. Pardini M, Brown JWL, Magliozzi R, Reynolds R, Chard DT. Surface-in pathology in multiple sclerosis: a new view on pathogenesis? Brain. 2021;144(6):1646–54.

33. Kochanski RB, Dawe R, Kocak M, Sani S. Identification of Stria Medullaris Fibers in the Massa Intermedia Using Diffusion Tensor Imaging. World Neurosurg. 2018;112:e497–e504.

34. Umapathy L, Keerthivasan MB, Zahr NM, Bilgin A, Saranathan M. Convolutional Neural Network Based Frameworks for Fast Automatic Segmentation of Thalamic Nuclei from Native and Synthesized Contrast Structural MRI. Neuroinformatics. 2022;20(3):651–64.

35. Morell-Ortega S, Ruiz-Perez M, Gadea M, Vivo-Hernando R, Rubio G, Aparici F, et al. Robust deep MRI contrast synthesis using a prior-based and task-oriented 3D network. Imaging Neurosci (Camb). 2025;3.

