## supplementary-material for "Interthalamic adhesion alterations in multiple sclerosis: associations with thalamic damage and cognition"

**Neuropsychological tests**

Information processing speed was assessed using five measures: the number of correct responses on the Computerized Speed and Capacity Test (CSCT), alertness subtests from the Test for Attentional Performance (TAP) (with and without an auditory warning cue), and reaction times during visual scanning in the TAP (with and without a target). Impairment in this domain was defined as a Z-score <-1.5 in at least two out of these five tests.

Attention was evaluated based on the number of errors on the CSCT and the number of correct responses on the TAP visual scanning task (with and without a target). A Z-score <-1.5 on any one of these tests indicated attentional impairment.

Working memory was assessed using the Paced Auditory Serial Addition Test (PASAT) (total number of correct responses) and the backward digit span task (reverse span length). Impairment was defined by a Z-score <-1.5 in either of the two tests.

Verbal memory was evaluated using the California Verbal Learning Test (CVLT), considering both the total score across the five immediate recall trials and the long-delay free recall score. Impairment was defined as a Z-score <-1.5 on either metric.

Visuospatial memory was assessed using the Brief Visuospatial Memory Test - Revised (BVMT-R), focusing on the total score across the first three immediate recall trials and the delayed recall score. Impairment was defined as a Z-score <-1.5 on either metric.

Executive functioning was evaluated using phonemic and semantic verbal fluency tasks. The domain was considered impaired if at least one of these tests yielded an abnormal result.

**Parameters of MRI acquisitions**

All participants underwent brain MRI on a 3T scanner (Vantage Galan 3T/ZGO; Canon Medical Systems) equipped with a 32-channel phased-array head coil. The parameters of the sequences were as follow:

(i) 3D gradient echo T1-weighted sequence: TR/TE/TI/flip angle = 2500 / 2.8 /950 ms / 9°; resolution: 1×1×1 mm³; field of view: 180 x 224 x 224 mm^3^,
(ii) 3D fluid-attenuated inversion recovery (FLAIR) sequence: TR/TE/TI = 7000 / 445.5 / 2100 ms; resolution: 1×1×1 mm³; FOV: 180 x 224 x 224 mm^3^,
(iii) 3D white matter-nulled magnetization-prepared rapid gradient echo (WMn-MPRAGE) sequence, previously optimized to enhance visualization of internal thalamic anatomy and MS-related thalamic lesions: TS/TR/TE/TI/flip angle = 4500 / 7.8 / 3.6 / 470 ms / 7°; resolution: 1×1×1 mm³; FOV: 180 x 224 x 180 mm^3^.

**Automatic delineation of IA**

The network architecture was a 3DUNET variant trained using pytorch 2.4 with a loss based on dice index and binary cross-entropy. The input consisted of both the standard 3D-T1 and the 3D-WMn images, while the output was the binary mask representing the IA from all the patients and healthy controls. The automated pipeline included the following steps:

(i) denoising of the input images (1),
(ii) N4 bias field correction to reduce intensity inhomogeneity (2),
(iii) registration to MNI space using ANTs (3),
(iv) intensity normalization (4) and standardization (z-scoring) of both the T1 and WMn images,
(v) cropping around the IA region,
(vi) application of super-resolution (factor ×2) using a residual CNN (5),
(vii) execution of the segmentation via the trained CNN,
(viii) and reintegration of the predicted mask into the full image space by reversing the cropping step.

A T1-only version was also developed which followed the same pipeline with the inclusion of a WMn synthesis step to compute the missing WMn image using a recent deep leaning 3D synthesis method (6). This pipeline will be included in the upcoming version of DeepThalamus 2.0 (7). The full pipeline including thalamic nuclei segmentation and intherthalamic adhesion has a running time of 90 seconds approximately.

**Supplementary figures**

**Supplementary Figure 1.**

**Performance of automatic IA delineation**

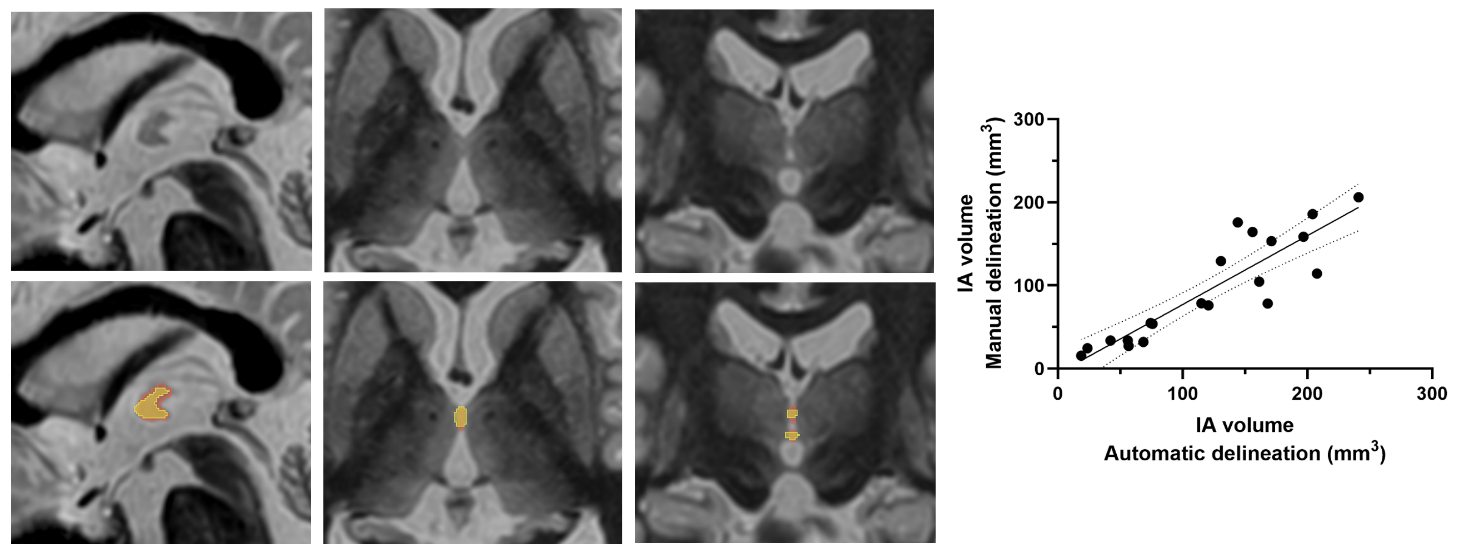
White matter-nulled images in sagittal, axial, and coronal planes from the external validation dataset showing a bilobar IA. Manual delineation is indicated by red contours, and automatic delineation by yellow contours. The scatter plot shows the correlation between manual and automatic IA volumes, with the linear regression line and its 95% confidence interval.

**Supplementary Figure 2.**

**Subtypes of IA**

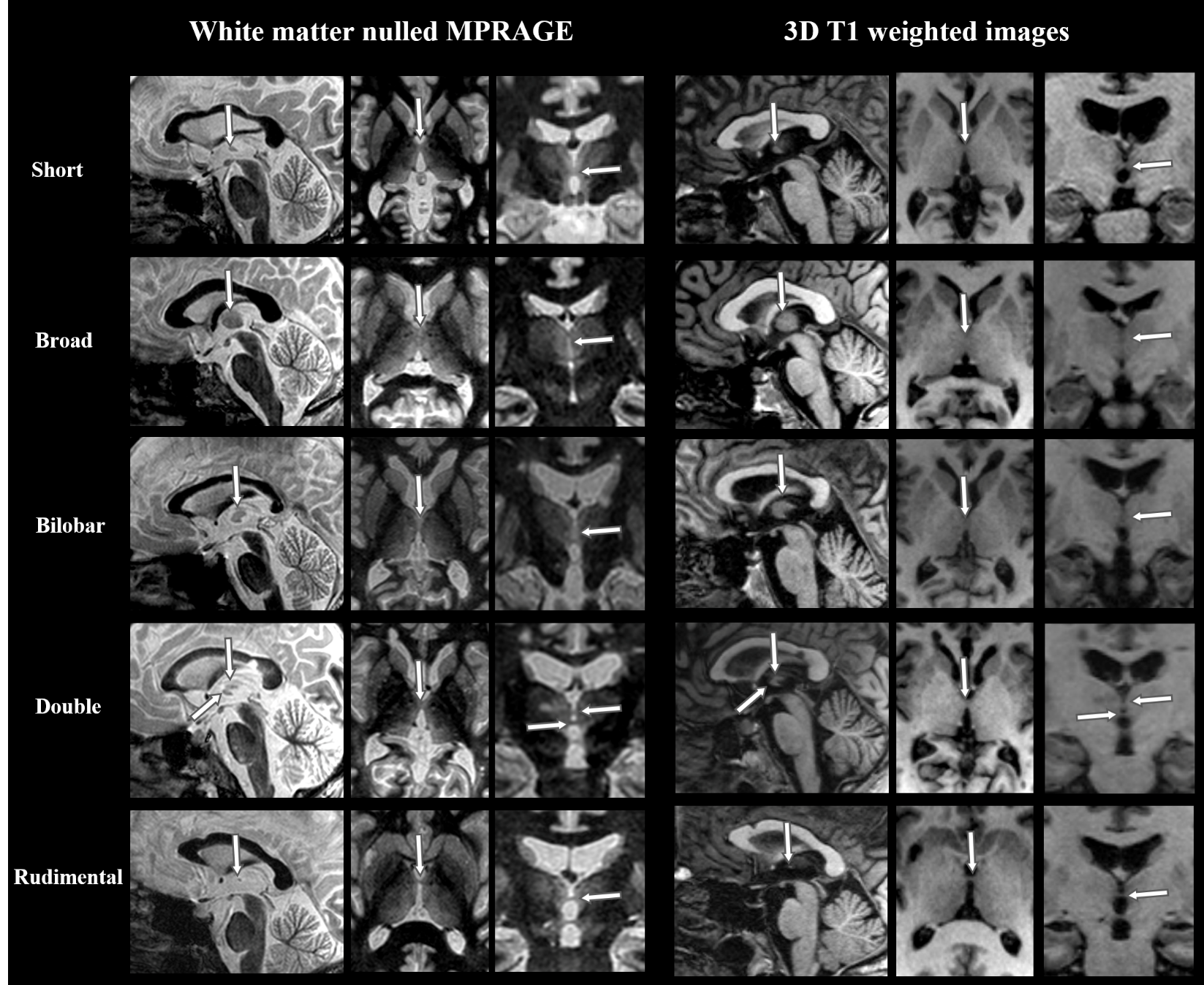
Representative examples of the five IA subtypes shown in orthogonal planes using either white matter-nulled (WMn) MPRAGE or standard 3D T1-weighted sequences.

**Supplementary Figure 3.**

**Appearance of IA on WMn Images**

Illustration of two broad (**A** and **B**) and one short (**C**) IA. Sagittal views are shown both with and without annotations, with IAs manually delineated in red. Red arrows indicate the stria medullaris, and red arrowheads point to the anterior commissures. Blue dotted lines mark the positions of the axial slices, while yellow arrowheads indicate the anterior and posterior boundaries of the IA.

The signal of the IA is not completely nulled by comparison with a pure white matter structure such as the anterior commissure. In the broad subtypes, a signal intensity gradient is visible, with lower signal in the antero-inferior portion, suggesting a higher fiber content (*), gradually increasing toward the postero-superior region.

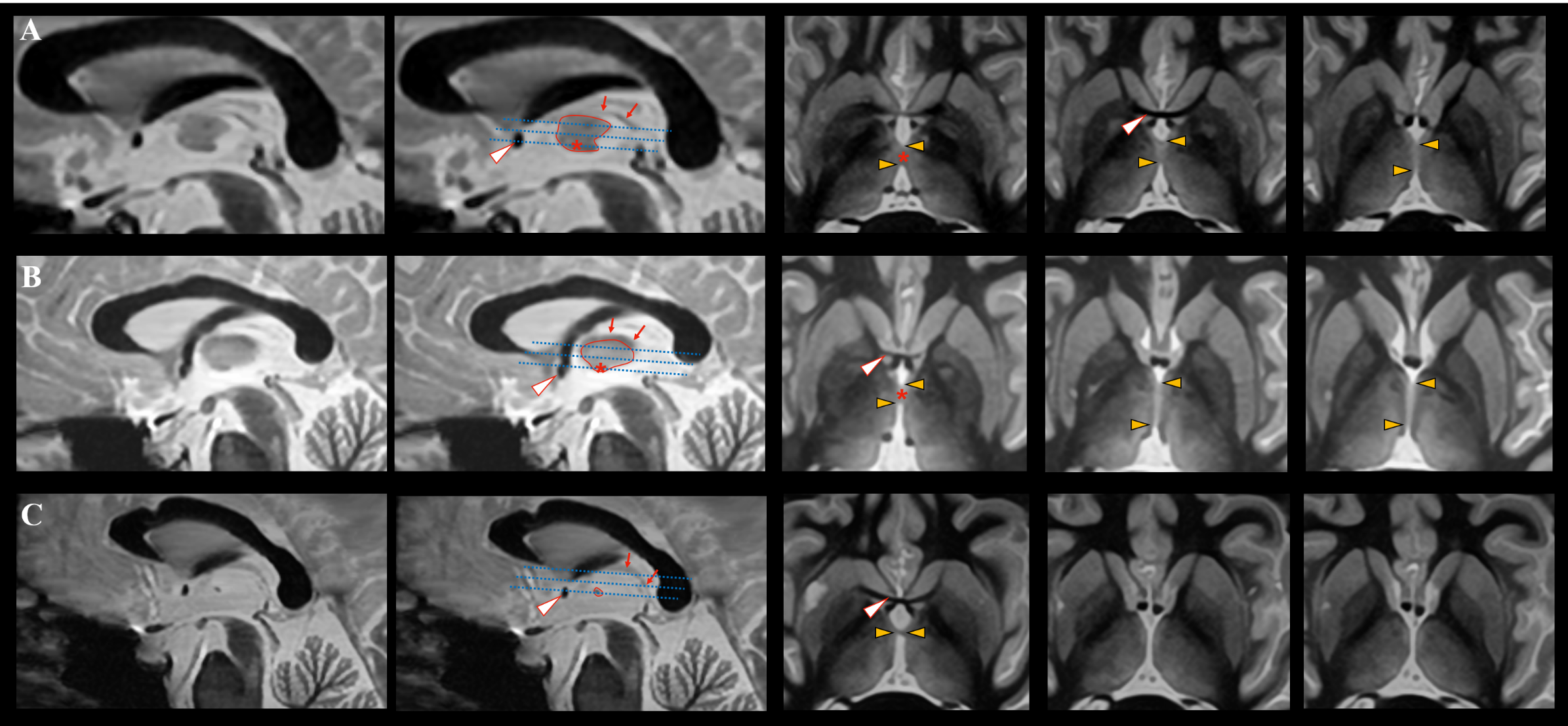

**Supplementary Figure 4.**

**IA length and height**

IA length and height (mm) are plotted in healthy controls (HC) and patients as well as according to the MS phenotype. CIS, clinically isolated syndrome, RRMS, relapsing remitting multiple sclerosis, PPMS, primary progressive multiple sclerosis.

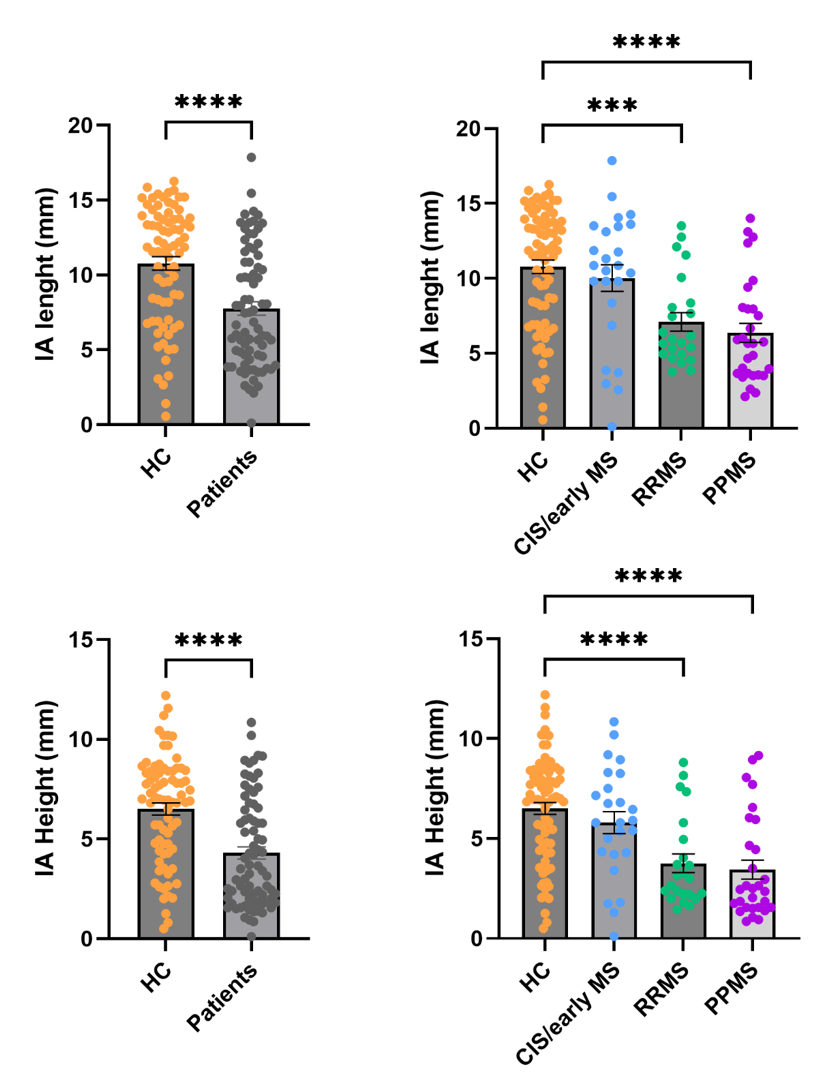

**Supplementary figure 5.**

**Individual Z-scores to the cognitive tests**

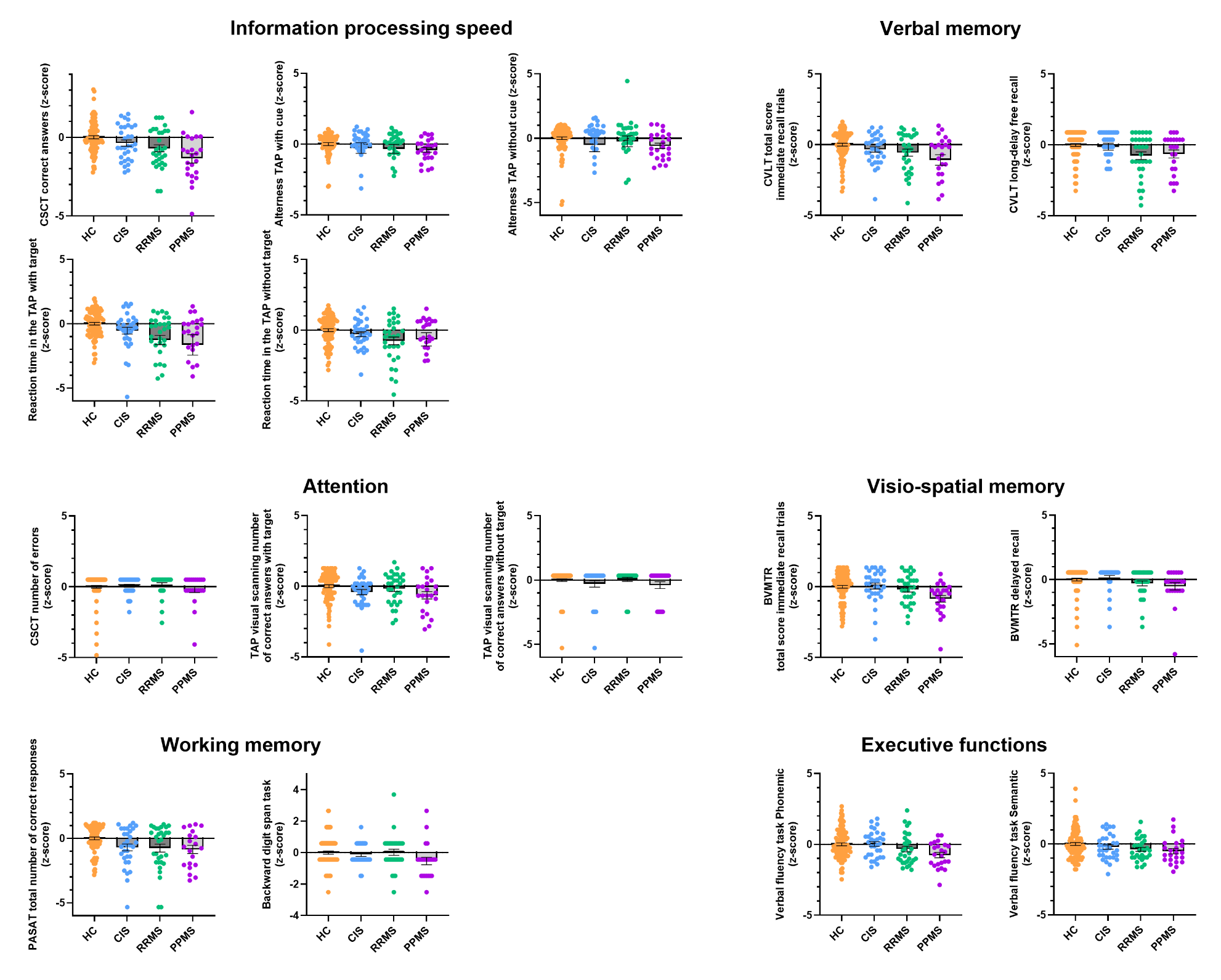

**Supplementary references**

1.	Manjon JV, Coupe P, Marti-Bonmati L, Collins DL, Robles M. Adaptive non-local means denoising of MR images with spatially varying noise levels. J Magn Reson Imaging. 2010;31(1):192-203.

2.	Tustison NJ, Avants BB, Cook PA, Zheng Y, Egan A, Yushkevich PA, et al. N4ITK: improved N3 bias correction. IEEE Trans Med Imaging. 2010;29(6):1310-20.

3.	Avants BB, Tustison NJ, Song G, Cook PA, Klein A, Gee JC. A reproducible evaluation of ANTs similarity metric performance in brain image registration. Neuroimage. 2011;54(3):2033-44.

4.	Manjon JV, Tohka J, Garcia-Marti G, Carbonell-Caballero J, Lull JJ, Marti-Bonmati L, et al. Robust MRI brain tissue parameter estimation by multistage outlier rejection. Magn Reson Med. 2008;59(4):866-73.

5.	Morell-Ortega S, Ruiz-Perez M, Gadea M, Vivo-Hernando R, Rubio G, Aparici F, et al. DeepCERES: A deep learning method for cerebellar lobule segmentation using ultra-high resolution multimodal MRI. Neuroimage. 2025;308:121063.

6.	Morell-Ortega S, Ruiz-Perez M, Gadea M, Vivo-Hernando R, Rubio G, Aparici F, et al. Robust deep MRI contrast synthesis using a prior-based and task-oriented 3D network. Imaging Neurosci (Camb). 2025;3.

7.	Ruiz-Perez M, Morell-Ortega S, Gadea M, Vivo-Hernando R, Rubio G, Aparici-Robles F, et al. DeepThalamus: A novel deep learning method for automatic segmentation of brain thalamic nuclei from multimodal ultra-high resolution MRI. arXiv. 2025.
